# Is sleep apnoea a risk factor for Covid-19? Findings from a retrospective cohort study

**DOI:** 10.1101/2020.05.14.20098319

**Authors:** Thijs Feuth, Tarja Saaresranta, Antti Karlsson, Mika Valtonen, Ville Peltola, Esa Rintala, Jarmo Oksi

## Abstract

**Background:** In the early phase of the coronavirus disease-19 (Covid-19) pandemic, Southwest Finland remained relatively spared. By the 3^rd^ of May 2020, a total of 28 patients have been admitted to the Turku University Hospital. In this paper, we explore baseline characteristics in order to identify risk for severe Covid-19 disease and critical care admission.

**Methods:** For this retrospective cohort study, data were derived from hospital records. Basic descriptive statistics were used to characterise patients, including medians, percentiles and frequencies. Differences were tested with Mann Whitney U-test and Pearson’s chi-square test.

**Results:** Pre-existent obstructive sleep apnoea (OSA) was present in 29% of patients admitted in the hospital for Covid-19, none of them having severe OSA. Overall, other findings on admission were comparable with those reported elsewhere. C-reactive protein (CRP) and procalcitonin (PCT) were higher in patients who were eventually transferred to critical care in comparison to in those who were not (median CRP 187 mg/L versus 52 mg/L, p<0.005 and median PCT 0.46 versus 0.12, p=0.047). Moreover, there was a trend towards lower oxygen saturation on admission in ICU-patients (87% versus 93%, p=0.09).

**Discussion:** OSA was pre-existent in a disproportional large group of patients, which suggests that it is an important risk factor for severe Covid-19. Furthermore, we identified high CRP, PCT and possibly oxygen saturation as useful clinical measures to identify patients at risk for critical care.

## Introduction

As the newly discovered Severe acute respiratory syndrome coronavirus 2 (SARS-CoV-2) is rapidly spreading around the world, Finland outside its capital Helsinki remains relatively spared in the early phase of the coronavirus disease-19 (Covid-19) pandemic. On the 3^rd^ of May 2020, WHO reported 3,349,786 cases worldwide, of which 1,518,895 in Europe alone.^1^ By then, Southwest Finland, home to 479,234 people, had a reported incidence of Covid-19 of 263 cases. Of them, 28 patients had been admitted to the Turku University Hospital.

In the first phase of the outbreak, cough and fever were the most reported symptoms.^2^ Advanced age, cardiovascular diseases and hypertension were among the first risk factors for severe disease and mortality to be recognized.^3,4^ In addition, obesity predisposes to severe Covid-19 requiring admission to critical care.^5^ Several mechanisms have been proposed to explain the link between obesity, cardiovascular disease and Covid-19, including low-grade chronic inflammation, chronic hypoxemia and oxidative stress and involvement of the renin angiotensin aldosterone system (RAAS) linked to expression of ACE2, the cellular receptor of SARS-CoV-2. However, the contribution of these mechanisms to Covid-19 pathology remain to be resolved.^6-9^

In this article, we report on the cases admitted to the Turku University Hospital, Finland, in the early phase of the pandemic.

## Methods

In April 2020, we started an observational database to systematically collect clinical data of all Covid-19 patients admitted in the University Hospital of Turku. In this report, we document on all cases who were admitted by May 3^rd^ 2020. Data cut-off was May 7^th^.

Baseline clinical findings are first clinical measurements as reported upon admission at the emergency room. In cases with no newly measured body weight or height measured during admission, older values were retracted from the patient files if available. Baseline laboratory tests were taken directly on admission or on the next weekday. For the subanalysis of obstructive sleep apnoea (OSA) patients, baseline sleep apnoea data was retracted from patient files and from the ResMed CPAP-telemonitoring system.

In all patients, SARS-CoV-2 was tested by polymerase chain reaction (PCR).

### Statistics

For descriptive statistics, median and 25^th^ to 75^th^ percentiles and percentages in case of frequencies were reported. Differences in continuous variables between ICU- and non-ICU patients were tested with the Mann-Whitney U test. Pearson’s chi-squared test was used to test for differences in frequencies.

### Ethics statement

According to the Finnish Medical Research Act (488/1999, sections 1-3), non-interventional studies do not require separate approval of the ethics committee. Approval for this study was obtained from the Turku University Hospital Clinical Research Centre, who did not require ethics committee approval or informed consent. The research was conducted according to the principles of the World Medical Association Declaration of Helsinki.

## Results

By May 3^rd^ 2020, a total of 28 patients had been admitted to the Turku University Hospital, Finland. By then, a total of 278 patients were diagnosed with Covid-19 by public services out of a total population of 479,234 people. Baseline clinical findings are summarized in table 1. Of note, female patients made up to 46% of admitted cases. The cohort includes only 1 minor of 15 years old. A total of 7 patients (25%) were treated on ICU. By the 10^th^ of May, 2 patients included in this study were still admitted in the hospital, with 1 on ICU. Median length of stay in hospital was 9.5 days, and median length of stay in ICU was 19 days. Fifteen patients (54%) could be discharged to home, 9 (32%) were discharged to a regional health centre. By May 10^th^, 2 patients had died in hospital, and 1 after referral to a regional health centre.

**Table 1:**
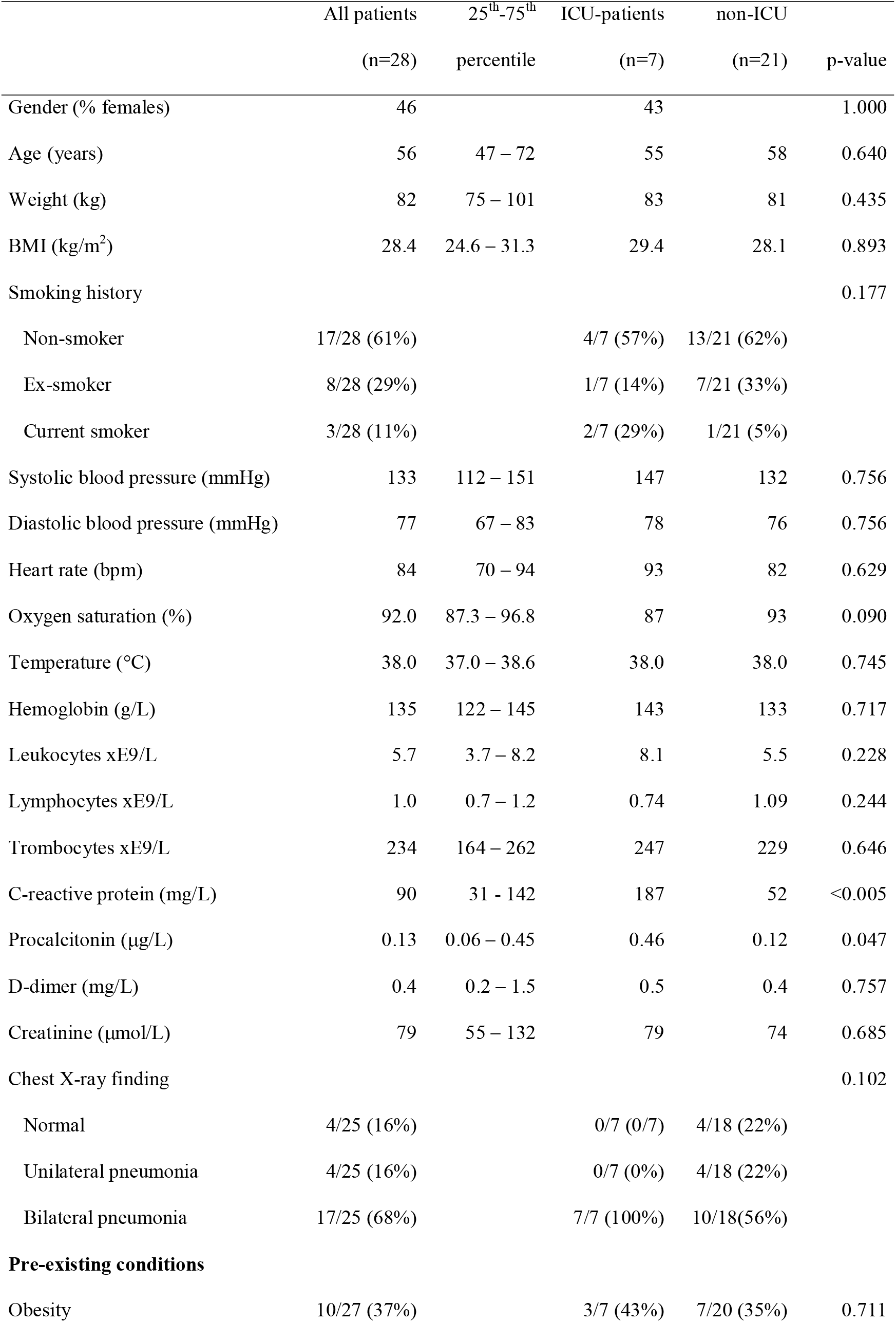
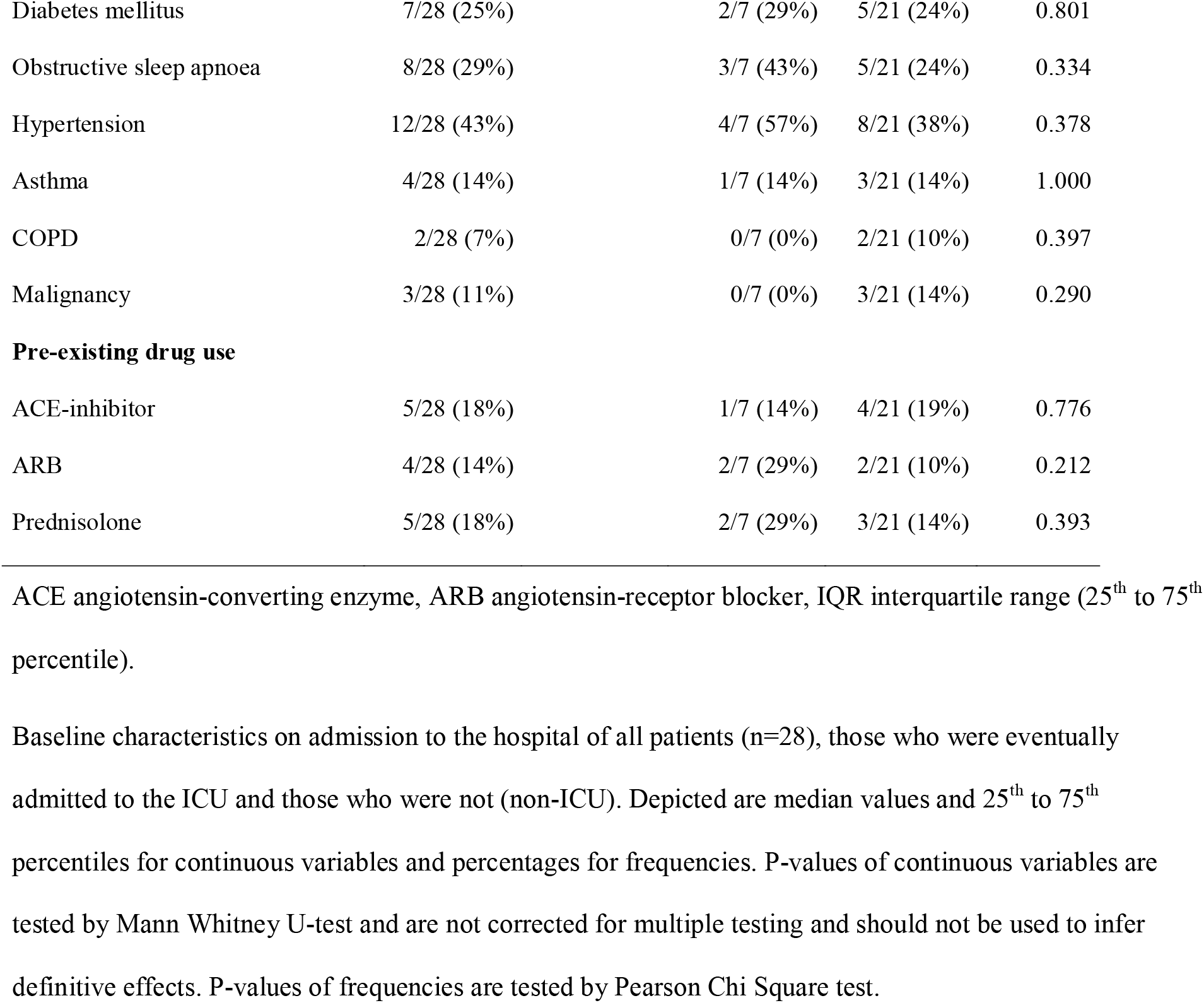
Baseline patient characteristics

|  | All patients<br>(n=28) | 25<sup>th</sup>-75<sup>th</sup><br>percentile | ICU-patients<br>(n=7) | non-ICU<br>(n=21) | p-value |
| --- | --- | --- | --- | --- | --- |
| Gender (% females) | 46 |  | 43 |  | 1.000 |
| Age (years) | 56 | 47 – 72 | 55 | 58 | 0.640 |
| Weight (kg) | 82 | 75 – 101 | 83 | 81 | 0.435 |
| BMI (kg/m<sup>2</sup>) | 28.4 | 24.6 – 31.3 | 29.4 | 28.1 | 0.893 |
| Smoking history |  |  |  |  | 0.177 |
| Non-smoker | 17/28 (61%) |  | 4/7 (57%) | 13/21 (62%) |  |
| Ex-smoker | 8/28 (29%) |  | 1/7 (14%) | 7/21 (33%) |  |
| Current smoker | 3/28 (11%) |  | 2/7 (29%) | 1/21 (5%) |  |
| Systolic blood pressure (mmHg) | 133 | 112 – 151 | 147 | 132 | 0.756 |
| Diastolic blood pressure (mmHg) | 77 | 67 – 83 | 78 | 76 | 0.756 |
| Heart rate (bpm) | 84 | 70 – 94 | 93 | 82 | 0.629 |
| Oxygen saturation (%) | 92.0 | 87.3 – 96.8 | 87 | 93 | 0.090 |
| Temperature (°C) | 38.0 | 37.0 – 38.6 | 38.0 | 38.0 | 0.745 |
| Hemoglobin (g/L) | 135 | 122 – 145 | 143 | 133 | 0.717 |
| Leukocytes xE9/L | 5.7 | 3.7 – 8.2 | 8.1 | 5.5 | 0.228 |
| Lymphocytes xE9/L | 1.0 | 0.7 – 1.2 | 0.74 | 1.09 | 0.244 |
| Trombocytes xE9/L | 234 | 164 – 262 | 247 | 229 | 0.646 |
| C-reactive protein (mg/L) | 90 | 31 - 142 | 187 | 52 | <0.005 |
| Procalcitonin (µg/L) | 0.13 | 0.06 – 0.45 | 0.46 | 0.12 | 0.047 |
| D-dimer (mg/L) | 0.4 | 0.2 – 1.5 | 0.5 | 0.4 | 0.757 |
| Creatinine (µmol/L) | 79 | 55 – 132 | 79 | 74 | 0.685 |
| Chest X-ray finding |  |  |  |  | 0.102 |
| Normal | 4/25 (16%) |  | 0/7 (0/7) | 4/18 (22%) |  |
| Unilateral pneumonia | 4/25 (16%) |  | 0/7 (0%) | 4/18 (22%) |  |
| Bilateral pneumonia | 17/25 (68%) |  | 7/7 (100%) | 10/18(56%) |  |
| **Pre-existing conditions** |  |  |  |  |  |
| Obesity | 10/27 (37%) |  | 3/7 (43%) | 7/20 (35%) | 0.711 |

|  |  |  |  |  |
| --- | --- | --- | --- | --- |
| Diabetes mellitus | 7/28 (25%) | 2/7 (29%) | 5/21 (24%) | 0.801 |
| Obstructive sleep apnoea | 8/28 (29%) | 3/7 (43%) | 5/21 (24%) | 0.334 |
| Hypertension | 12/28 (43%) | 4/7 (57%) | 8/21 (38%) | 0.378 |
| Asthma | 4/28 (14%) | 1/7 (14%) | 3/21 (14%) | 1.000 |
| COPD | 2/28 (7%) | 0/7 (0%) | 2/21 (10%) | 0.397 |
| Malignancy | 3/28 (11%) | 0/7 (0%) | 3/21 (14%) | 0.290 |
| **Pre-existing drug use** |  |  |  |  |
| ACE-inhibitor | 5/28 (18%) | 1/7 (14%) | 4/21 (19%) | 0.776 |
| ARB | 4/28 (14%) | 2/7 (29%) | 2/21 (10%) | 0.212 |
| Prednisolone | 5/28 (18%) | 2/7 (29%) | 3/21 (14%) | 0.393 |

ACE angiotensin-converting enzyme, ARB angiotensin-receptor blocker, IQR interquartile range (25<sup>th</sup> to 75<sup>th</sup> percentile).
Baseline characteristics on admission to the hospital of all patients (n=28), those who were eventually admitted to the ICU and those who were not (non-ICU). Depicted are median values and 25<sup>th</sup> to 75<sup>th</sup> percentiles for continuous variables and percentages for frequencies. P-values of continuous variables are tested by Mann Whitney U-test and are not corrected for multiple testing and should not be used to infer definitive effects. P-values of frequencies are tested by Pearson Chi Square test.

Of patients of whom smoking status was known, 11% were current smokers and 29% ex-smokers. Median blood pressure (133/77 mmHg) and heart rate (84 /min) were in the normal range, as were hemoglobin (135 g/L), thrombocytes (234 e9/L), leukocytes (5.7 e9/L), and creatinine (79 μmol/L). Fever (median 38.0°C), hypoxemia (median SaO_2_ 92%), lymphocytopenia (median 1.0 xe9/L), increased C-reactive protein (CRP, median 90 mg/L) and increased procalcitonin (PCT, median 0.13 μg/L) were common. Chest X-ray (CXR) showed bilateral opacities in 68% of cases and unilateral opacities in 16%. Common pre-existent conditions included hypertension in 43% of cases, obesity (in 35%), OSA (29%), diabetes mellitus (25%), asthma (14%), COPD (7%) and active malignant disease (11%). Baseline characteristics are summarized in table 1.

Baseline characteristics were compared between patients who were later on admitted to ICU (n=7) and those who were not (non-ICU, n=21). Baseline CRP (median 187 mg/L) and PCT (median 0.46 μg/L) were significantly higher among those who were later on transferred to ICU compared to those who were not (median CRP 52 mg/L, p<0.005 and median PCT 0.12, p=0.047). In addition, a trend towards lower SaO_2_ on admission was observed in ICU-patients (87%) in comparison with non-ICU patients (93%, p=0.09). All ICU-patients had bilateral opacities in CXR on admission to the hospital (table 1 and figure 1).

**Figure 1:**
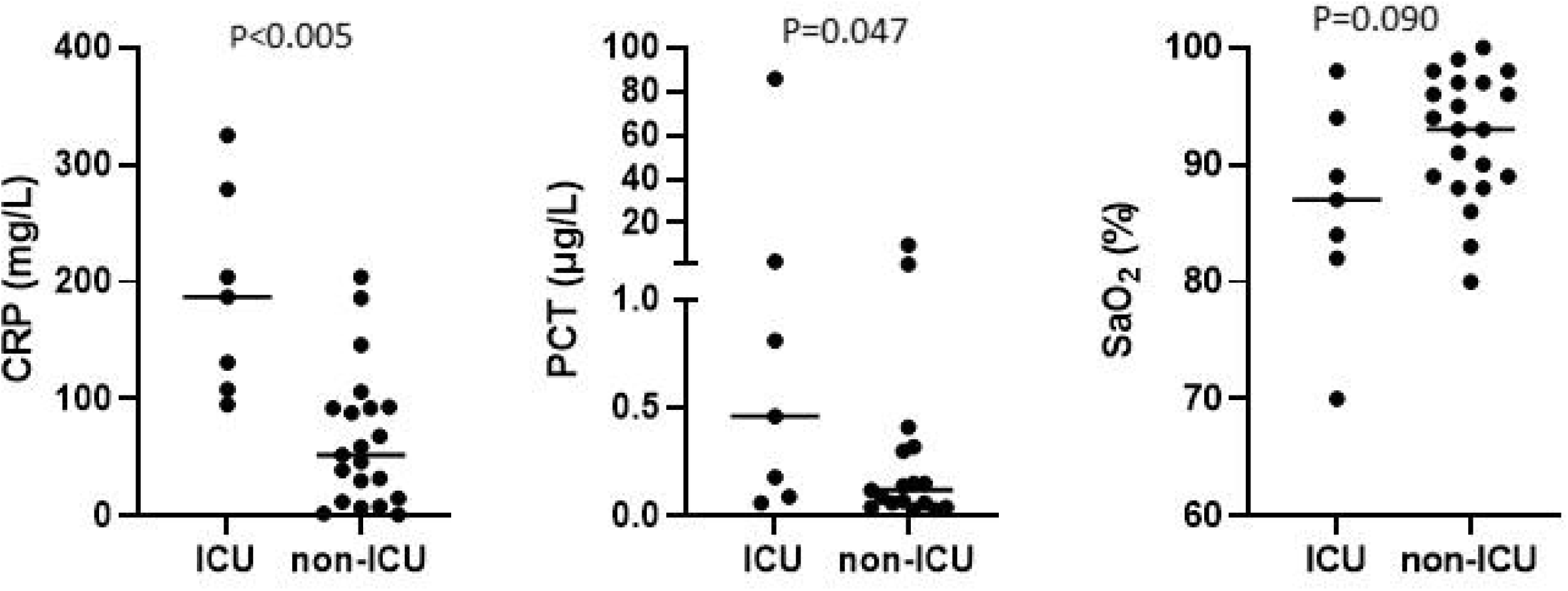
Baseline characteristics and admission to critical care CRP C-reactive protein. PCT procalcitonin, SaO_2_ oxygen saturation. CRP, procalcitonin and SaO_2_ on admission in patients who later on required admission to the ICU and those who were treated on conventional ward (non-ICU). Horizontal lines represents median values.

In the Hospital District of Southwest Finland, a total of 12,799 people (2.7%) are treated with continuous positive airway pressure (CPAP) therapy and around 2,000 people with mandibular advancement device (MAD) for OSA. Thus, prevalence of hospital-treated OSA is around 3.1%.

As prevalence of previously diagnosed OSA was disproportionally high in this cohort, we performed a post-hoc descriptive analysis of those patients. BMI >30 kg/m^2^ was found in 75% of OSA patients. Median BMI was 38.0 kg/m^2^. Thus, the majority of OSA patients in this cohort were severely obese. Median time of diagnosis of OSA was 2.5 years before Covid-19 and at time of diagnosis, median apnoea-hypopnoea index (AHI) was 18 /h. Median 3% oxygen desaturation index (ODI3) was 12 /h. At time of diagnosis of OSA, average overnight SpO_2_ was 92.1% and minimum was 83.5% (median values) and morning pCO_2_ values were in the normal range in all cases (median 4.9 kPa). One patient was using MAD and the other 7 patients were CPAP-users (six using automatic positive airway pressure (APAP) device and one fixed pressure CPAP). CPAP adherence was good (median 7.5 hours) in those actually using the device (n=5), but two patients had discontinued before admission for Covid-19. Characteristics of OSA patients in our case series are summarized in table 2.

**Table 2:**
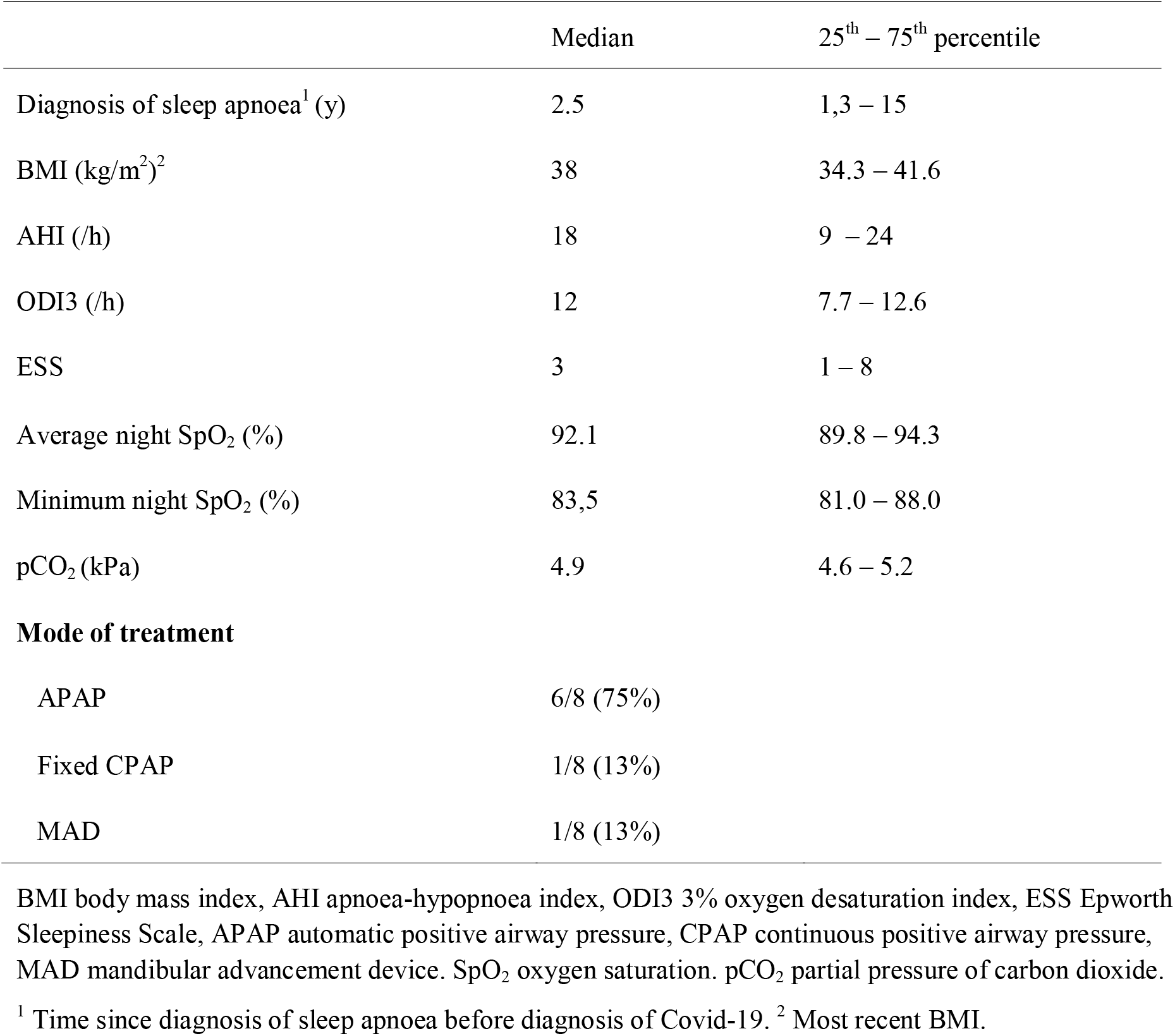
Characteristics of pre-existent sleep apnoea in patients with Covid-19

|  | Median | 25<sup>th</sup> – 75<sup>th</sup> percentile |
| --- | --- | --- |
| Diagnosis of sleep apnoea<sup>1</sup> (y) | 2.5 | 1,3 – 15 |
| BMI (kg/m<sup>2</sup>)<sup>2</sup> | 38 | 34.3 – 41.6 |
| AHI (/h) | 18 | 9 – 24 |
| ODI3 (/h) | 12 | 7.7 – 12.6 |
| ESS | 3 | 1 – 8 |
| Average night SpO<sub>2</sub> (%) | 92.1 | 89.8 – 94.3 |
| Minimum night SpO<sub>2</sub> (%) | 83,5 | 81.0 – 88.0 |
| pCO<sub>2</sub> (kPa) | 4.9 | 4.6 – 5.2 |
| **Mode of treatment** |  |  |
| APAP | 6/8 (75%) |  |
| Fixed CPAP | 1/8 (13%) |  |
| MAD | 1/8 (13%) |  |

BMI body mass index, AHI apnoea-hypopnoea index, ODI3 3% oxygen desaturation index, ESS Epworth Sleepiness Scale, APAP automatic positive airway pressure, CPAP continuous positive airway pressure, MAD mandibular advancement device. SpO<sub>2</sub> oxygen saturation. pCO<sub>2</sub> partial pressure of carbon dioxide.
<sup>1</sup> Time since diagnosis of sleep apnoea before diagnosis of Covid-19. <sup>2</sup> Most recent BMI.

## Discussion

Most of our findings were generally consistent with reports of Covid-19 cases elsewhere.^2,4,5^ However, two of our findings are of special interest. Pre-existent OSA was present in 29% of cases being over ten times higher than the prevalence of OSA patients under treatment in our region. Because of this disproportionally high frequency of OSA, we performed a post-hoc analysis of those patients and their pre-existent condition. Even though obesity is by now an established risk factor for severe Covid-19, weight alone does not explain the high proportion of patients with OSA, as obesity is a much more common pre-existing condition than OSA in the Finnish population with a prevalence of 26.1% among men and 27.5% among women.^10^ In fact, OSA was present in 21% of cases in a case series in Seattle and in 28.6% of critically ill Covid-19 patients in a cohort of Washington.^11,12^ Several mechanisms may contribute to increased risk of severe Covid-19 in OSA patients, independent of overweight. Intermittent hypoxemia is one of the main pathologic features of OSA and could potentially worsen hypoxemia caused by Covid-19.^13^ OSA is also related to chronic inflammatory state and is associated with increased levels of proinflammatory markers such as ferritin, IL-6 and leptin, which may contribute to the risk for cytokine storm in Covid-19.^13-15^ Hypercoagulability is another pathophysiologic feature shared by both Covid-19 and OSA.^16,17^ Furthermore, RAAS may also be influenced by sleep apnoea, and RAAS-associated regulation of ACE2-expression could be another mechanism by which OSA may contribute to the risk of severe Covid-19.^7,8,18^

A second striking observation is the high level of CRP in patients admitted to the ICU. Even though CRP has a well-established role in diagnostics and follow-up of bacterial pneumonia and other bacterial infections, the usefulness of CRP levels has been less clear. Even though several studies were not able to distinguish viral pneumonia from a pneumonia of bacterial origin based on CRP, CRP is often being used as a guidance for treatment with antibiotics.^19^ Our data shows that high CRP is associated with increased risk for ICU admission within several days. CRP levels may reflect the pulmonary damage caused by Covid-19. Theoretically, concomitant bacterial pneumonia can be present but undiagnosed. However, we found no evidence of bacterial infection, blood cultures remained negative in all patients and those with CRP >100 mg/L upon presentation had in most cases radiology findings consistent with Covid-19 without radiologic suspicion of bacterial pneumonia.

According to our findings, high CRP and PCT and possibly low SaO_2_ on admission may be predictors for transfer to critical care. If confirmed in larger studies, this may be useful in triage which may be especially helpful when the epidemic is out of control and hospital places are scarce, requiring decision making based on basic clinical and laboratory assessment. The usability of CRP, PCT and SaO_2_ in clinical workup of Covid-19 has yet to be confirmed in larger studies.

The high prevalence of pre-existing OSA in this cohort may have implications for individual risk assessment and may help to understand pathogenesis of severe Covid-19. However, the finding should be considered with extreme caution, as this cohort is rather small. The question whether OSA is an independent risk factor should be addressed in larger cohorts.

Our study has some major limitations. First, the size of the study remains small due to low incidence of Covid-19 in Southwest Finland in the early phase of the pandemic. Second, due to the retrospective nature of this study, there was incomplete documentation on some of the variables. Third, follow-up was very short, which may lead to under reporting of mortality and complications, although the number still in hospital on the date of data cut-off was very low. Fourth, we only report on patients admitted in the hospital, which leads to an obvious bias, mild cases are largely missed, and fragile patients with severe disease may not be transported to the hospital but receive instead palliative care at home.

## Data Availability

The data that support the findings of this study are available on request from the corresponding author, TF. The data are not publicly available due to their containing information that could compromise the privacy of research participants.

## Acknowledgement

We are grateful to Ton Feuth for his advices on the statistics used in this study.

## Notes

### Competing Interest Statement

The authors have declared no competing interest.

### Funding Statement

No external funding was received for this study.

## References

1. WHO. Coronavirus disease (COVID-19) Situation Report. In. Vol 104. May 3rd 20202020.

2. Guan WJ, Ni ZY, Hu Y, et al. Clinical Characteristics of Coronavirus Disease 2019 in China. N Engl J Med. 2020;382(18): 1708–1720.

3. Rúan Q, Yang K, Wang W, Jiang L, Song J. Clinical predictors of mortality due to COVID-19 based on an analysis of data of 150 patients from Wuhan, China. Intensive Care Med. 2020.

4. Mehra MR, Desai SS, Kuy S, Henry TD, Patel AN. Cardiovascular Disease, Drug Therapy, and Mortality in Covid-19. N Engl J Med. 2020.

5. Lighter J, Phillips M, Hochman S, et al. Obesity in patients younger than 60 years is a risk factor for Covid-19 hospital admission. Clin Infect Dis. 2020.

6. Sattar N, McInnes IB, McMurray JJV. Obesity a Risk Factor for Severe COVID-19 Infection: Multiple Potential Mechanisms. Circulation. 2020.

7. South AM, Diz DI, Chappell MC. COVID-19, ACE2, and the cardiovascular consequences. Am J Physiol Heart Circ Physiol. 2020;318(5):H1084–H1090.

8. Barceló A, Elorza MA, Barbé F, Santos C, Mayoralas LR, Agusti AG. Angiotensin converting enzyme in patients with sleep apnoea syndrome: plasma activity and gene polymorphisms. Eur Respir J. 2001;17(4):728–732.

9. Pazarlı AC, Ekiz T, ilik F. Coronavirus disease 2019 and obstructive sleep apnea syndrome. Sleep Breath. 2020.

10. Koponen P, Borodulin K, Lundqvist A, Sääksjärvi K, Koskinen S. Terveys, toimintakyky ja hyvinvointi Suomessa FinTerveys 2017-tutkimus. In: THL.

11. Bhatraju PK, Ghassemieh BJ, Nichols M, et al. Covid-19 in Critically Ill Patients in the Seattle Region - Case Series. N Engl J Med. 2020.

12. Arentz M, Yim E, Klaff L, et al. Characteristics and Outcomes of 21 Critically Ill Patients With COVID-19 in Washington State. JAMA. 2020.

13. McSharry D, Malhotra A. Potential influences of obstructive sleep apnea and obesity on COVID-19 severity. J Clin Sleep Med. 2020.

14. Kheirandish-Gozal L, Gozal D. Obstructive Sleep Apnea and Inflammation: Proof of Concept Based on Two Illustrative Cytokines. Int J Mol Sci. 2019;20(3).

15. Ip MS, Lam KS, Ho C, Tsang KW, Lam W. Serum leptin and vascular risk factors in obstructive sleep apnea. Chest. 2000;118(3):580–586.

16. Connors JM, Levy JH. Thromboinflammation and the hypercoagulability of COVID-19. J Thromb Haemost. 2020.

17. Alonso-Fernández A, Toledo-Pons N, García-Río F. Hypercoagulability, Obstructive Sleep Apnea, and Pulmonary Embolism. JAMA Otolaryngol Head Neck Surg. 2018;144(5):459.

18. Jin ZN, Wei YX. Meta-analysis of effects of obstructive sleep apnea on the renin-angiotensin-aldosterone system. J Geriatr Cardiol. 2016;13(4):333–343.

19. Thomas J, Pociute A, Kevalas R, Malinauskas M, Jankauskaite L. Blood biomarkers differentiating viral versus bacterial pneumonia aetiology: a literature review. Ital J Pediatr. 2020;46(1):4.

